# Neurofilament light and glial fibrillary acidic protein in mood and anxiety disorders: A systematic review and meta-analysis

**DOI:** 10.1101/2024.03.07.24303938

**Authors:** Matthew JY Kang, Jasleen Grewal, Dhamidhu Eratne, Charles Malpas, Wei-Hsuan Chiu, Kasper Katisko, Eino Solje, Alexander F Santillo, Philip B. Mitchell, Malcolm Hopwood, Dennis Velakoulis

## Abstract

**Background:** Neurofilament light chain (NfL) and glial fibrillary acidic protein (GFAP) are biomarkers of neuronal injury measurable in cerebrospinal fluid (CSF) and blood. Despite their potential as diagnostic tests for neurodegenerative disorders, it is unclear how they behave in mood and anxiety disorders. We conducted a systematic review and meta-analysis to investigate whether NfL and GFAP concentrations were altered in adults with mood and anxiety disorders compared to healthy controls.

**Methods:** The study was prospectively registered on PROSPERO (CRD42023434617). We followed the PRISMA guidelines, searched PubMed, Web of Science, PsycINFO, MEDLINE and Embase up to the 31/05/2023, and assessed relevant studies and their risk of bias. The primary outcome was the standardised mean difference (SMD) and 95% confidence interval (95% CI) of NfL and GFAP concentrations, which was pooled using a random-effects model adopting the restricted maximum likelihood estimator.

**Results:** Twenty-one studies met inclusion criteria, comprising of 2327 individuals (695 major depression, 502 bipolar disorder, and 1130 controls). When we compared people with major depression and controls, there was no difference in NfL (SMD = 0.29; 95% CI: -0.10, 0.68) nor GFAP (SMD = 0.47; 95% CI: -0.74, 1.68). In people with bipolar disorder, NfL was significantly elevated compared to controls (SMD = 0.58; 95% CI: 0.16, 0.99). However, the subgroup analysis including more sensitive assay kits (blood Simoa and CSF ELISA), found no significant difference (SMD = 0.40; 95% CI: -0.04, 0.85). Only one study studied GFAP in bipolar disorder. No studies explored NfL nor GFAP concentrations in anxiety disorders.

**Discussion:** We found that NfL and GFAP concentrations were not elevated in depression. In bipolar disorder, NfL concentration was elevated, though not in the sensitivity analysis. Our study informs clinicians about how to interpret these emerging biomarkers in determining whether a person’s symptoms are caused by a neurodegenerative or mood disorder.

## Introduction

Recent developments in blood and cerebrospinal fluid (CSF) biomarkers in neurological conditions have offered exciting prospects for enhancing diagnostic assessments. Neurofilament light chain (NfL), a cytoskeletal protein of neurons, has emerged as a sensitive biomarker for assessing neuronal injury (1). Neurodegenerative processes lead to the release of NfL into the CSF, which subsequently pass through to the bloodstream at lower concentrations (2). Although initial studies were limited to measuring NfL in CSF, technological advances have facilitated precise quantification of NfL in blood (3). In particular, blood NfL measured using single-molecule array digital immunoassay (Simoa) correlate well with CSF NfL (4). Numerous studies have shown that NfL, in both CSF and blood, is a valuable marker for diagnosing, assessing disease severity, and predicting prognosis in a diverse range of neurodegenerative conditions, including Alzheimer’s disease and frontotemporal lobar degeneration (5–7). NfL can also distinguish neurodegenerative disorders from primary psychiatric disorders within memory clinics and neuropsychiatry services (8–10). More recently, glial fibrillary acidic protein (GFAP), a component of glial cell cytoskeleton, has emerged as a sensitive measure of astrogliosis and neuroinflammation, demonstrating superior prognostic accuracy to NfL in multiple sclerosis (11,12).

Given the potential clinical utility of these biomarkers in determining whether an individual’s undifferentiated presentation with psychiatric, cognitive and/or neurological symptoms is due to a neurological or a psychiatric cause, it is crucial to understand how NfL or GFAP behave in primary mood or anxiety disorders. If NfL and GFAP levels remain within normal ranges in primary mood and anxiety disorders, which are the two most prevalent mental disorders (13), such findings would support the use of these biomarkers in distinguishing neurodegenerative conditions at diagnostic assessments. Conversely, should they be elevated in primary mood and anxiety disorders, this may provide insights into their neurobiological underpinnings.

One of the earliest investigations into NfL and GFAP in psychiatric disorders involved a cohort of elderly women who remained dementia-free during a 10-year follow-up period (14). Those with major depression exhibited significantly higher concentrations of baseline CSF NfL compared to the rest of the group (major depression mean: 427 ± 186 vs control mean: 277 ± 186 ng/L), whilst there was no difference in the GFAP concentrations (major depression mean: 946 ± 196 vs control: 887 ± 308 ng/L). Subsequent studies comparing blood NfL and GFAP in mood disorders to healthy controls have yielded mixed findings, either finding elevated concentrations in mood disorders (15–17), or no difference (6,18).

More recent studies have examined the relationship between NfL or GFAP with clinical outcomes in mood disorders. The concentration of GFAP was higher in people with major depression and correlated with the severity of depressive symptoms (17). NfL was found to be negatively correlated with cognitive processing speed in people with major depression compared to healthy controls (19). Several theories have been proposed to explain why NfL or GFAP may be elevated in mood disorders, including astroglia dysfunction (17), vascular pathology (14), and neuroinflammatory processes (20).

Given these conflicting findings in this area of major clinical and scientific importance, we have undertaken a systematic review and meta-analysis of studies investigating NfL and GFAP concentrations in people with mood (depression and bipolar disorder) and anxiety disorders compared to healthy controls. Secondly, we sought to identify any associations between the severity of mood and anxiety disorders with NfL and GFAP.

## Methods

This systematic review was developed using the Preferred Reporting Items for Systematic Reviews and Meta-Analysis (PRISMA) guidelines (21). The protocol was prospectively registered with PROSPERO (CRD42023434617).

### Eligibility criteria

All eligible studies met the following criteria: (1) adult patients diagnosed with either mood (depression and bipolar disorder) or anxiety disorder according to DSM or ICD (22–25); and (2) measured NfL or GFAP in plasma, serum or CSF measured by Simoa, electrochemiluminescence method (ECL) or enzyme linked immunosorbent assay (ELISA).

Case-series, meeting abstracts, reviews and meta-analyses were excluded. For articles with overlapped samples, only the report with largest sample size was included.

Exclusions criteria were: (1) mood or anxiety disorders due to general medical conditions including dementia and stroke, and (2) co-morbid neurological disorders including multiple sclerosis and dementia.

### Search strategy

Electronic searches were conducted *via* the Web of Science, PubMed, MEDLINE(OVID), Embase and PsycINFO from the inception of the databases through to 31st May 2023. The search terms included (“neurofilament light*” OR “NfL”) AND (“glial fibrillary acidic protein*” OR “GFAP”) AND (“depressi*” OR ”bipolar*” OR “anxiet*” OR “mood disorder*”). Studies were required to be in English. We have described our full search strategy in the supplementary file.

### Study Selection and Data Collection

We used Cochrane’s Covidence, a web-based systematic review manager, for study selection and quality analysis. One investigator (MK) screened the title and abstracts. Two investigators (MK and JG) independently reviewed the main reports and supplementary materials for their eligibility. We resolved discrepancies by consensus and involved a third senior investigator (MH) when required.

The primary outcome was the mean difference of NfL and GFAP in people with mood and anxiety disorders. The mean and standard deviation (SD) of NfL and GFAP was collected in the different cohorts. If this was not available, authors were contacted for the data. If the authors did not respond, the following methods were used to convert the data according to Cochrane Handbook guidelines (26): 1) median, interquartile ranges and minimum/maximum values were converted using an estimation formula (27,28), 2) mean and SD of subgroups (i.e. male and female) were combined using Cochrane’s formula (29), and 3) studies that only presented the results in plots were estimated using WebPlotDigitzer (30).

Our secondary outcomes were as follows: 1) mood state and severity (measured by a standardised rating scale), 2) duration of mood/anxiety disorder, and 3) cognitive impairment (measured by a standardised rating scale). We also collected known covariates of NfL and GFAP if they were reported, including age and weight (31).

### Quality of individual studies

Two investigators (MK and JG) independently assessed the quality of included studies using the Newcastle-Ottawa Scale (NOS) quality assessment tool (32). NOS contains a total of 8 small items, and evaluates three aspects: selection, comparability and outcome. Studies were classified as “poor quality” (0-2 points), “average quality” (3-5 points), and “good or high quality” (6-9 points).

### Statistical analysis

All statistical analyses were performed using R version 4.2.2 (2022-10-31) and the meta package (33). Meta-analyses were separately conducted for individual psychiatric diagnoses (i.e. major depression and bipolar disorder). For each diagnosis, we calculated standardised mean differences (SMDs) of each biomarker (NfL and GFAP) between psychiatric disorders and controls to be able to pool the different types of assay kits. Of note, Random effects meta-analysis models, rather than fixed effects models, were selected a priori based on the theoretical absence of a consistent ‘true effect’ across the studies. The restricted maximum likelihood estimator was adopted. Ninety-five percent confidence intervals (95% CI) and prediction intervals (PI) were calculated and visualised using forest plots. A PI quantifies the likely range in which a new study’s effect size will fall into, assuming that this future study is of a similar nature to those meta-analysed. We assumed a two-sided *P* < 0.05 to indicate statistical significance.

Where there were a sufficient number of studies, sensitivity analyses were performed to ensure any findings were consistent with the main analyses: type of biomarker analysis (i.e. CSF ELISA and serum/plasma Simoa; blood Simoa) and studies that used age-matched controls. We intended to analyse further subgroups including duration of psychiatric illness and age at diagnosis, however there was an insufficient number of studies.

We performed meta regression on the severity of mood symptoms as measured by the scales used in the study (i.e. Hamilton’s Depression Rating Scale; HDRS and Montgomery-Asberg

Depression Rating Scale; MADRS). We could not perform other meta regression analyses including variables such as cognition, duration of psychiatric illness and number of mood disorder episodes due to the limited numbers of studies.

As a post-hoc analysis to include studies (all of which used Simoa assay kits) that did not have any control groups, we used a robust mixed-means model to compare the mean concentration of NfL and GFAP between cases (depression or bipolar) and controls.

We assessed for publication bias using funnel plots of the effect sizes of mood disorders for NfL and GFAP. We performed meta-analyses only where there were at least five studies to ensure acceptable statistical power to make an inference. I^2^ and tau^2^ statistics were used to quantify between-study heterogeneity in the meta-analyses.

## Results

### Study selection

The systematic search identified 2,890 unique studies, of which 110 were selected for full-text review. Twenty-one studies met our inclusion criteria for the meta-analysis, as shown in Figure 1. Twelve studies investigated major depression, five bipolar disorder, four reported data on both, and no studies reported on other types of mood disorders nor anxiety disorders.

**Figure 1.**
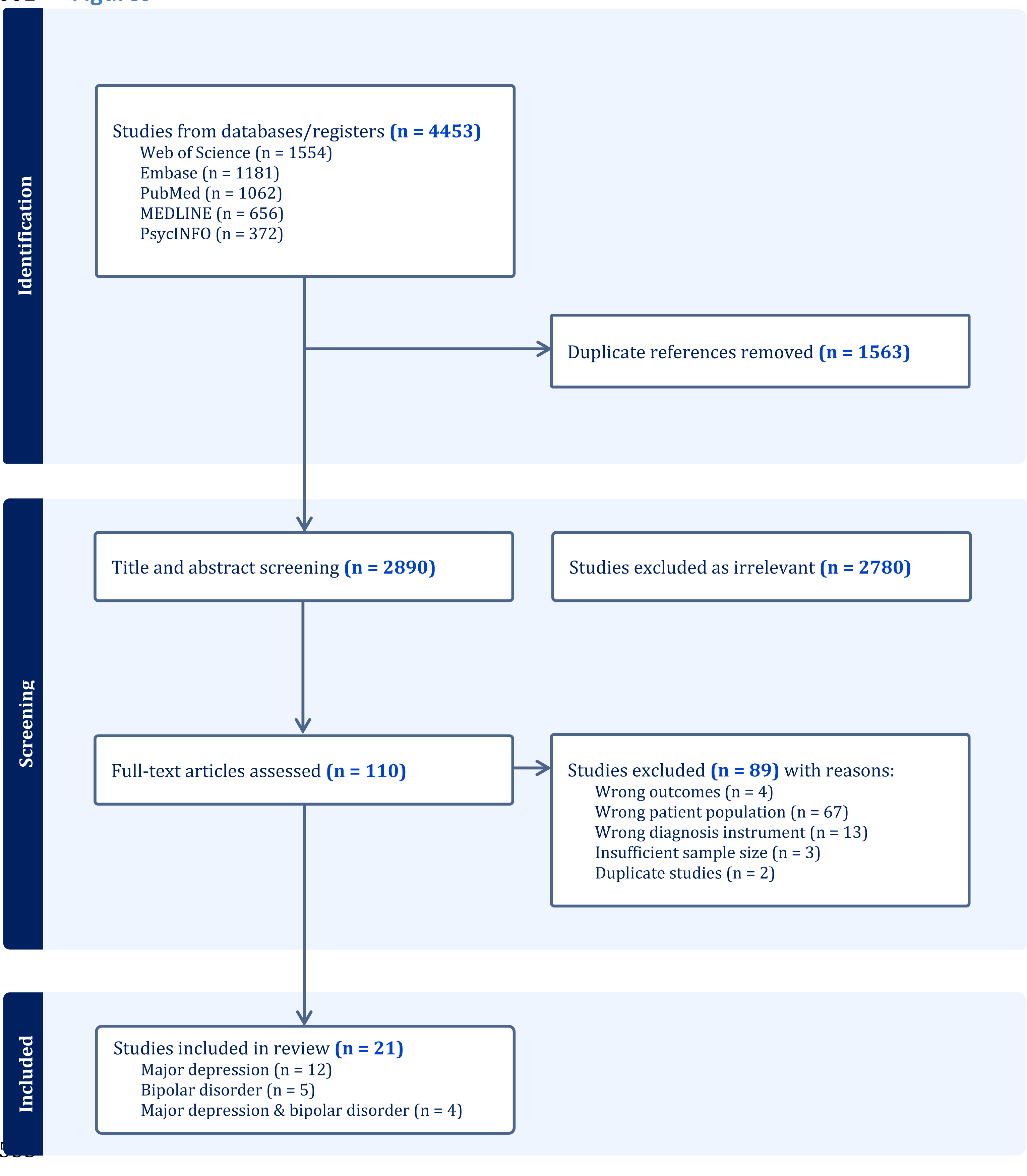
PRISMA Diagram for study selection

The main reason that some full-text articles were excluded was due to studies either not reporting or collecting individual psychiatric diagnoses. A number of studies used a less rigorous process to collect diagnoses (self-report, file review and diagnosis based on a cut-off score from a mood rating scale) (34). Four studies had an insufficient sample size (n<10) (9,35,36). Some studies used indirect measures of NfL and GFAP, including extracellular vesicles, mRNA, IgG autoantibodies and urine (37–40).

### Study characteristics

Final sample data were obtained for 2327 individuals, of which 695 had major depression, 502 bipolar disorder, and 1130 healthy controls. Two studies (41,42) did not have a healthy control group, so were only included in the additional mixed-model meta-analyses. Seventeen studies analysed blood samples, three used CSF, and Knorr et al’s study (16) investigated both CSF and blood. The majority of the studies (75%) were published after 2020, reflecting the relatively recent availability of Simoa and ELISA technology to measure NfL and GFAP in blood. (Tables 1-2)

**Table 1.** Characteristics of studies included: NfL in major depression.

| Authors and year | Country | Sample Type | Analysis type | Diagnosis classification | Mood state | Sample size |  | Age Mean (SD) |  | Sex (Female %) |  | NFL concentration^ |  | Total NOS Score |
| --- | --- | --- | --- | --- | --- | --- | --- | --- | --- | --- | --- | --- | --- | --- |
|  |  |  |  |  |  | Case | Control | Case | Control | Case | Control | Case | Control |  |
| Ashton et al, 2021 (6) | England | Plasma | Simoa | DSM-5 | Not specified | 51 | 114 | 32.3 (6.9) | 64.0 (14.1) | 69% | 55% | 11.19 (5.8) | 22.5 (10.7) | 3 |
| Bai et al, 2023 (43) | Taiwan | Serum | HRP ELISA | DSM-5 | HDRS ≤16 | 24 | 29 | 30.8 (13.0) | 30.9 (10.7) | 67% | 62% | 22.4 (12.3)** | 16.7 (6.8) | 5 |
| Bavato et al, 2021 (19) | Switzerland | Plasma | Simoa | DSM-5 | Clinically stable | 41 | 258 | 36.0 (10.9) | Unavailable | 59% | NA% | 26.3 (10.4) | 24.5 (9.6) | 5 |
| Besse et al, 2020 (44) | Germany | Serum | Simoa | ICD-10 | Moderate to severe depressive episode | 15 | 15 | 49.2 (14.0) | "± 3 years" | 73% | NA% | 17.4 (11.88) | Mean = 15.57<br>95% CI*: 10.69, 20.40 | 5 |
| Chen et al, 2022 (45) | Taiwan | Plasma | HRP ELISA | DSM-5 | Not specified | 40 | 40 | 28.3 (14.4) | 28.3 (14.1) | 68% | 68% | 28.8 (22.5)** | 16.7 (8.1) | 5 |
| Eratne et al, 2023 (8) | Australia | Plasma | Simoa | DSM-IV | Depressive phase ≥ moderate severity | 42 | 96 | 55.0 (12.9) | 44.5 (14.4) | 57% | 52% | 10.9 (7.0)** | 9.4 (14.2)** | 4 |
| Gudmundsson et al, 2010 (14) | Sweden | CSF | ELISA Sandwich | DSM-III-R | Not specified | 11 | 65 | Combined: 73.9 (3.2) |  | 100% | 100% | 427 (318) | 277 (186) | 5 |
| Huang et al, 2023 (18) | Taiwan | Serum | Simoa | DSM-IV-TR | Outpatients | 35 | 86 | 34.7 (10.3) | 33.4 (6.2) | 77% | 13% | 5.4 (2.2) | 6.7 (2.7) | 5 |
| Hviid et al, 2023 (15) | Denmark | Serum | Simoa | DSM-IV | Treatment naive | 110 | 33 | 42 (11) | 39 (14) | 65% | 70% | 7.7 (3.0)* | 7.6 (3.1)* | 5 |
| Jiang et al 2021, (46) | China | Serum | ELISA Sandwich | DSM-IV | First episode, HDRS > 16 | 82 | 72 | 34 (11) | 34 (12) | 61% | 49% | Median = 405.8<br>IQR: 281.5–625.5 | Median = 143.5<br>IQR: 73.6–339.3 | 5 |
| Katisko et al, 2020 (41) | Finland | Serum | Simoa | ICD-10 | Included moderate, severe & psychotic depression | 16 | 0 | 55.7 (6.7) | NA | 56% | NA | 12.4 (4.3)** | NA | 3 |
| Ramezani et al, 2022 (47) | Iran | Serum | ELISA | DSM-5 | Suicide attempt | 28 | 35 | M: 26.28 (9.37)<br>F: 26.61 (9.49) | M: 28.10 (8.54)<br>F: 30.76 (5.12) | 75% | 71% | M: 55.42 (33.63)<br>F: 36.66 (35.21) | M: 14.11 (4.38)<br>F: 13.57 (5.45) | 5 |
| Steinacker et al, 2021 (17) | Germany | Serum | Simoa | DSM-5 | Moderate-severe depressive episode | 45 | 16 | Median=48<br>Range 19-69 | Median=45<br>Range 27-64 | 64% | 75% | 29.3 (35.3) | 15.2 (7.1) | 4 |
| Wallensten et al, 2022 (48) | Sweden | Plasma | ELISA | ICD-10 | Not specified | 31 | 61 | 40.3 (10.8) | 42.9 (9.5) | 84% | 85% | 357.4 (87.1) | 306.4 (115.7) | 6 |

^CSF = ng/L, plasma/serum = pg/mL; \*estimates using WebPlotDigitizer; \*\*data obtained directly from corresponding author
CI = confidence interval, F = females, HDRS = Hamilton Depression Rating Scale, M = males, MADRS = Montgomery-Asberg Depression Rating Scale, NA = not applicable, NOS = Newcastle-Ottawa Scale, YMRS = Young Mania Rating Scale

**Table 2.** Characteristics of studies included: NfL in bipolar disorder.

| Authors and year | Country | Sample Type | Analysis type | Diagnosis classification | Mood state | Sample size |  | Age Mean (SD) |  | Sex (Female %) |  | NFL concentration^ |  | Total NOS Score |
| --- | --- | --- | --- | --- | --- | --- | --- | --- | --- | --- | --- | --- | --- | --- |
|  |  |  |  |  |  | Case | Control | Case | Control | Case | Control | Case | Control |  |
| Aggio et al, 2022 (49) | Italy | Plasma | Simoa | DSM-IV | Depressed | 45 | 29 | 48.2 (11.9) | 41.8 (10.2) | 78% | 55% | 9.1 (4.8) | 4.28 (2.39) | 3 |
| Al Shweiki et al, 2019 (50) | Germany | Serum | Simoa | DSM-5 | Manic, depressive and mixed states | 11 | 27 | Median = 51.4<br>IQR: 33.5-58.1 | Median = 46.8<br>IQR: 39.1-54.1 | 36% | 63% | Median = 17.8<br>IQR: 12.6 - 23.1 | Median = 17.8<br>IQR: 11.3 - 19.1 | 4 |
| Bai et al, 2023 (43) | Taiwan | Serum | HRP ELISA | DSM-5 | YMRS ≤12 | 25 | 29 | 33.2 (13.2) | 30.9 (10.7) | 60% | 62% | 28.5 (12.4)** | 16.7 (6.8) | 5 |
| Chen et al, 2022 (51) | Taiwan | Serum | Simoa | DSM-5 | Euthymic | 100 |  | 46.5 (11.7) | NA | 52% | NA% | 11.1 (8.1)** | NA | 5 |
| Eratne et al, 2023 (8) | Australia | Plasma | Simoa | DSM-IV | Depressive phase of at least moderate severity | 121 | 96 | 44.1 (12.18) | 44.48 (14.35) | 62% | 52% | 8.4 (4.8)** | 9.4 (14.2)** | 5 |
| Knorr et al, 2022 (16) | Denmark | CSF & Plasma | ELISA & Simoa | ICD-10 | In remission (YMRS ≤8; HDRS ≤8) | 85 | 44 | Median = 33<br>IQR: 26-42 | Median = 30<br>IQR: 25-42 | 51% | 52% | CSF: Median = 332<br>IQR: 246-479<br>Plasma: Median = 6.81<br>IQR: 4.97 - 9.07 | CSF: Median = 354<br>IQR: 214-566<br>Plasma: Median = 5.73<br>IQR: 4.50 - 7.84 | 6 |
| Ramezani et al, 2022 (47) | Iran | Serum | ELISA | DSM-5 | Suicide attempt | 22 | 35 | M: 33.00 (15.18)<br>F: 31.72 (13.10) | M: 28.10 (8.54)<br>F: 30.76 (5.12) | 82% | 71% | M: 46.50 (38.27)<br>F: 37.88 (31.63) | M: 14.11 (4.38)<br>F: 13.57 (5.45) | 5 |
| Rolstad et al, 2015 (52) | Sweden | CSF | ELISA | DSM-IV | Euthymic (MADRS < 14, YMRS < 14) | 82 | 71 | 38.3 (12.5) | 37.8 (14.6) | 59% | 62% | 485.7 (425.6) | 254.4 (55.4) | 5 |
| Steinacker et al, 2021 (17) | Germany | Serum | Simoa | DSM-5 | Manic, depressive and mixed states | 11 | 16 | Median = 48<br>Range: 19-69 | Median = 45<br>Range: 27-64 | 27% | 75% | 21.2 (16.6) | 15.2 (7.1) | 4 |

^CSF = ng/L, plasma/serum = pg/mL; \*estimates using WebPlotDigitizer; \*\*data obtained directly from corresponding author
CI = confidence interval, F = females, HDRS = Hamilton Depression Rating Scale, M = males, MADRS = Montgomery-Asberg Depression Rating Scale, NA = not applicable, NOS = Newcastle-Ottawa Scale, YMRS = Young Mania Rating Scale

**Table 3.** Characteristics of studies included: GFAP in major depression and bipolar disorder.

| Author | Country | Sample Type | Analysis type | Diagnosis classification | Mood state | Sample size |  | Age Mean (SD) |  | Sex (Female %) |  | GFAP concentration^ |  | Total NOS Score |
| --- | --- | --- | --- | --- | --- | --- | --- | --- | --- | --- | --- | --- | --- | --- |
|  |  |  |  |  |  | Case | Control | Case | Control | Case | Control | Case | Control |  |
| Major depressive disorder |  |  |  |  |  |  |  |  |  |  |  |  |  |  |
| Gudmundsson et al, 2010 (14) | Sweden | CSF | ELISA Sandwich | DSM-III-R | Not specified | 11 | 65 | Cases + controls: 73.9 (3.2) |  | 100% | 100% | 946 (196) | 887 (308) | 5 |
| Hviid et al, 2023 (15) | Denmark | Serum | Simoa | DSM-IV | Treatment naïve | 110 | 33 | 42 (11) | 39 (14) | 65% | 70% | 63.7 (23.8)* | 74.2 (35.2)* | 5 |
| Katisko et al, 2021 (42) | Finland | Serum | Simoa | ICD-10 | Included moderate, severe and psychotic depression | 22 | 0 | 55.7 (6.3) | NA | 55% | NA | 224 (113)** | NA | 3 |
| Michel et al, 2021 (20) | Germany | CSF | ELISA | ICD-10 | Inpatients | 102 | 39 | 44.2 (13.6) | 34.6 (12.0) | 54% | 85% | 733 (401) | 245.6 (176.3) | 5 |
| Steinacker et al, 2021 (17) | Germany | Serum | Simoa | DSM-5 | Moderate-severe depressive episode | 45 | 16 | Median = 48 Range 19-69 | Median = 45 Range 27-64 | 64% | 75% | 211 (99) | 147.0 (61.0) | 4 |
| Bipolar disorder |  |  |  |  |  |  |  |  |  |  |  |  |  |  |
| Steinacker et al, 2021 (17) | Germany | Serum | Simoa | DSM-5 | Manic, depressive and mixed states | 11 | 16 | Median = 48 Range 19-69 | Median = 45 Range 27-64 | 27% | 75% | 125 (50) | 147.0 (61.0) | 4 |

^CSF = ng/L, plasma/serum = pg/mL; \*estimates using WebPlotDigitizer; \*\*unpublished data obtained directly from corresponding author F = females, M = males, NA = not applicable, NOS = Newcastle-Ottawa Scale

Although Ashton et al. (2021) reported that the sample size of their control (cognitively unimpaired and CSF amyloid-β negative) and depression groups were 130 and 37 respectively, this conflicted with their supplementary data source table that reported 115 and 51 values respectively. We chose to perform the meta-analysis with the latter values, as they matched the sex proportions reported in their main table. Bavato et al. (2021) drew on 295 healthy controls and referenced their earlier work (Barro et al., 2018, n=258) for the characteristics of the healthy controls, which is included in this meta-analysis instead.

### Quality of studies

The quality of studies was considered at least moderate across studies. The area of low quality was in the domain of comparability due to the inequalities in age and sex between study groups, as summarised in the supplementary eTable 1.

### Synthesis of results

#### Major depression

Neurofilament light chain

***All studies (Figure 2a):*** There was no significant difference in the concentration of NfL in people with major depression compared to healthy controls (SMD = 0.29; 95% CI: -0.10, 0.68). The prediction interval for this pooled mean difference was very wide (PI: -1.11, 1.69) with significant heterogeneity (I^2^ = 90%; tau^2^ = 0.37). A regression-based Egger’s test did not show any evidence of small study effects (p-value = 0.35) though on visual inspection of the funnel plot (supplementary eFigure 1), Ashton et al’s study’s was an outlier (6). This study had a significant age difference between the depression and control groups (mean difference 31.7 years).

**Figure 2a.**
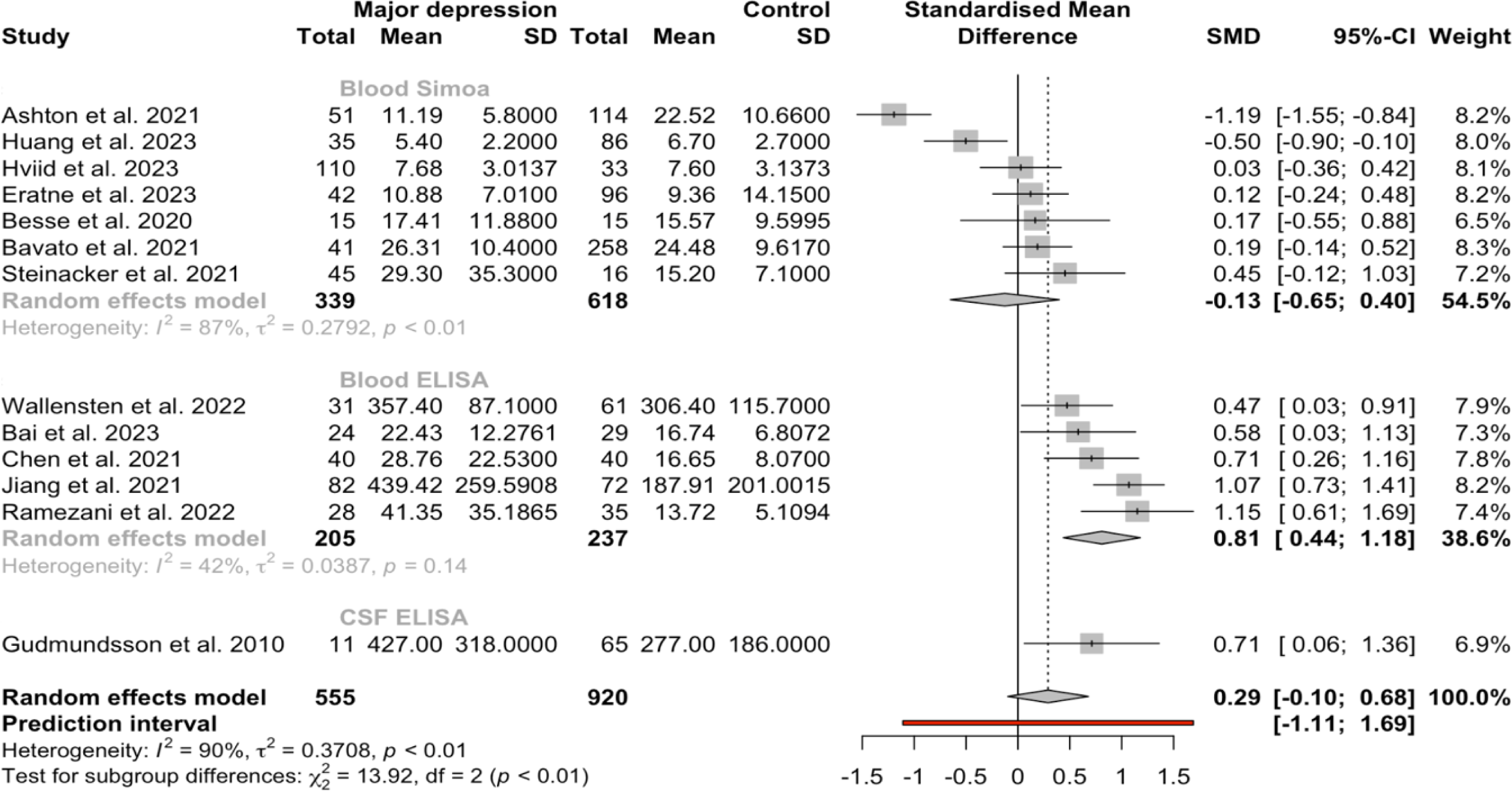
Forest plot of NfL in major depression (all studies)

*In the subgroup analysis of studies that included age-matched controls (Figure 2b),* NfL was significantly higher in major depression, though with a wide PI (SMD = 0.44; 95% CI: 0.11, 0.77; PI: -0.61, 1.50). *The subgroup analysis model which only included data using CSF ELISA and blood Simoa (Figure 2c)*, which are more accurate measures of NfL (2,4), found no significant difference (SMD = -0.03, 95% CI: -0.53, 0.47). The heterogeneity for both subgroup analysis models were high (I^2^ = 79%, tau^2^ = 0.17 and I^2^ = 57%, tau^2^ = 0.07 respectively).

**Figure 2b.**
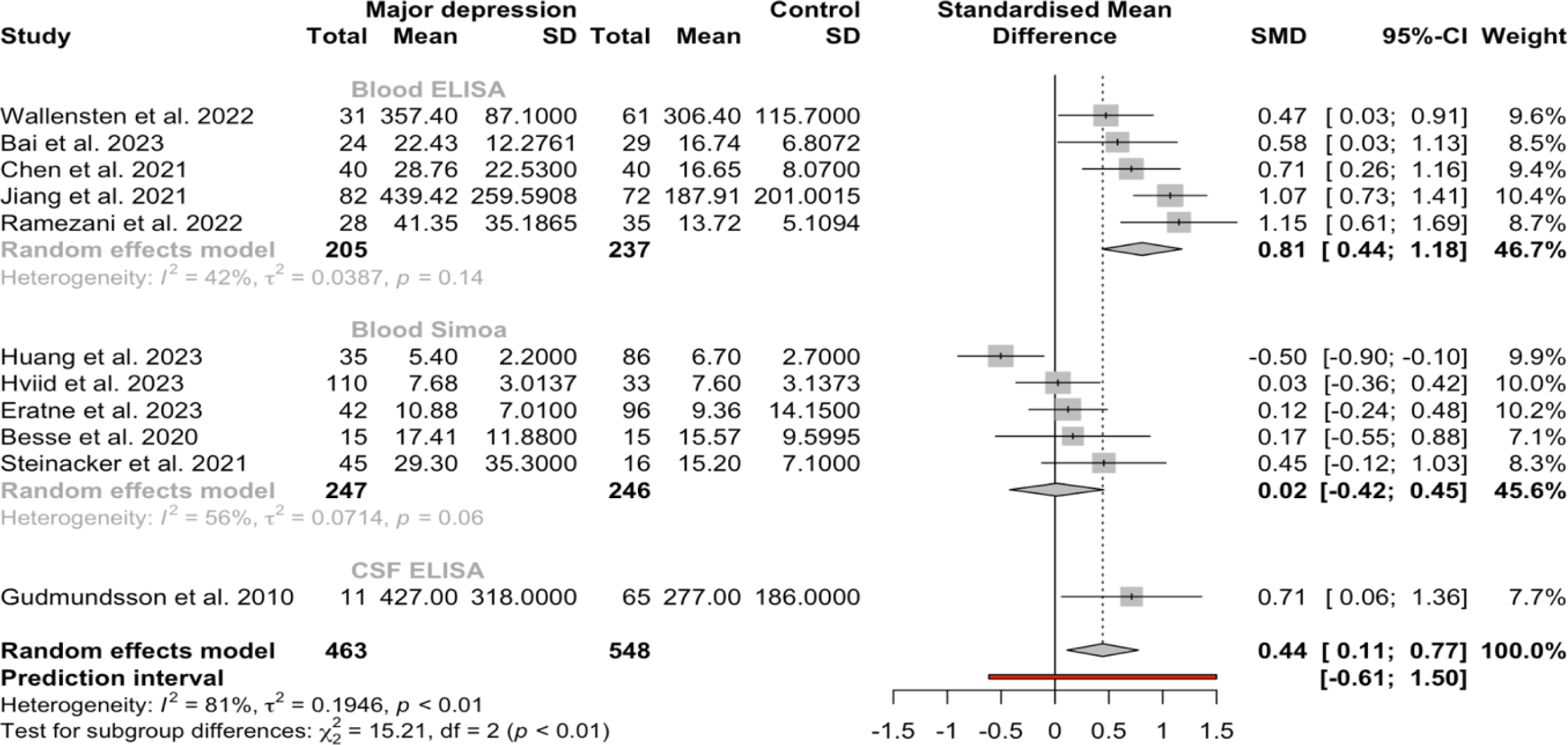
Forest plot of NfL in major depression (age-matched studies only)

Similarly, the subgroup analysis model including six age-matched studies that used CSF ELISA and Simoa also found no significant difference (SMD = 0.11 95% CI: -0.32, 0.55).

A random-effects meta-regression model which included all NfL studies suggested that mood severity did not influence NfL. In the subgroup mixed-model mean analysis of studies that only used blood Simoa, we found no significant difference in NfL levels.

Glial fibrillary acidic protein

GFAP (Figure 2d) was not significantly different between people with major depression and healthy controls (SMD = 0.47; 95% CI: -0.74, 1.68; PI: -3.09, 4.03), with significant heterogeneity between the studies (I^2^ = 92%, tau^2^ = 0.53). There was insufficient data to perform additional meta-regressions including cognition and GFAP.

**Figure 2c.**
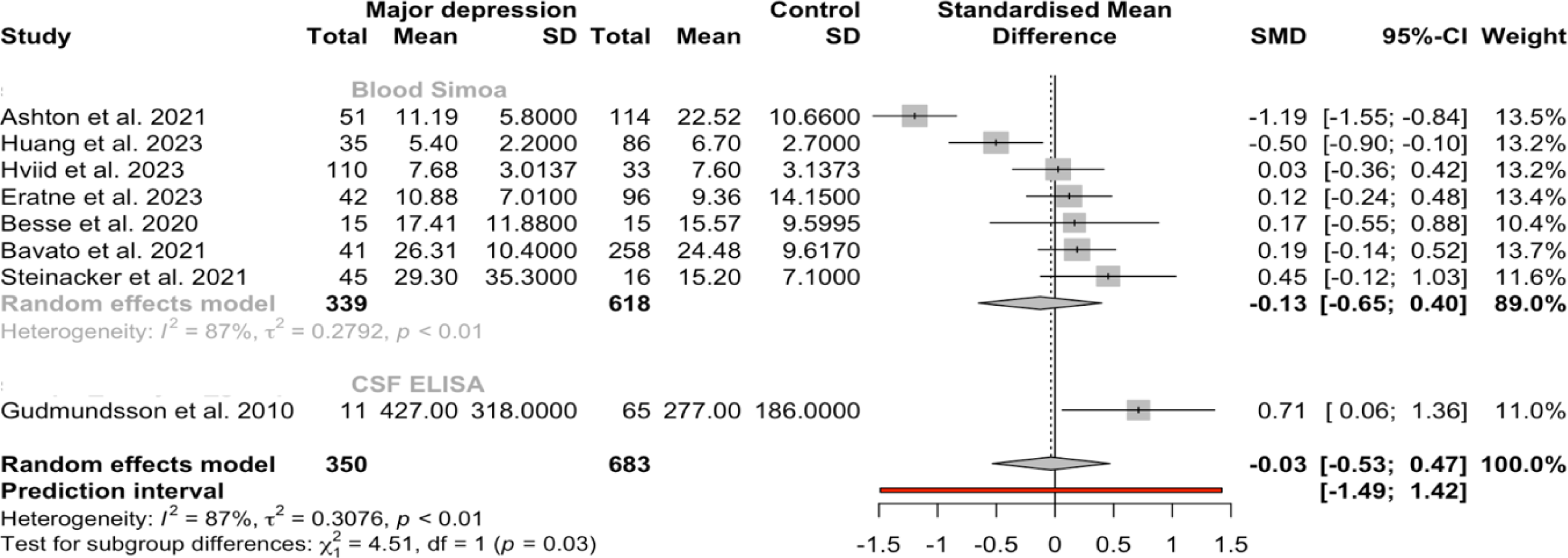
Forest plot of NfL in major depression (Simoa and CSF studies only)

**Figure 2d.**
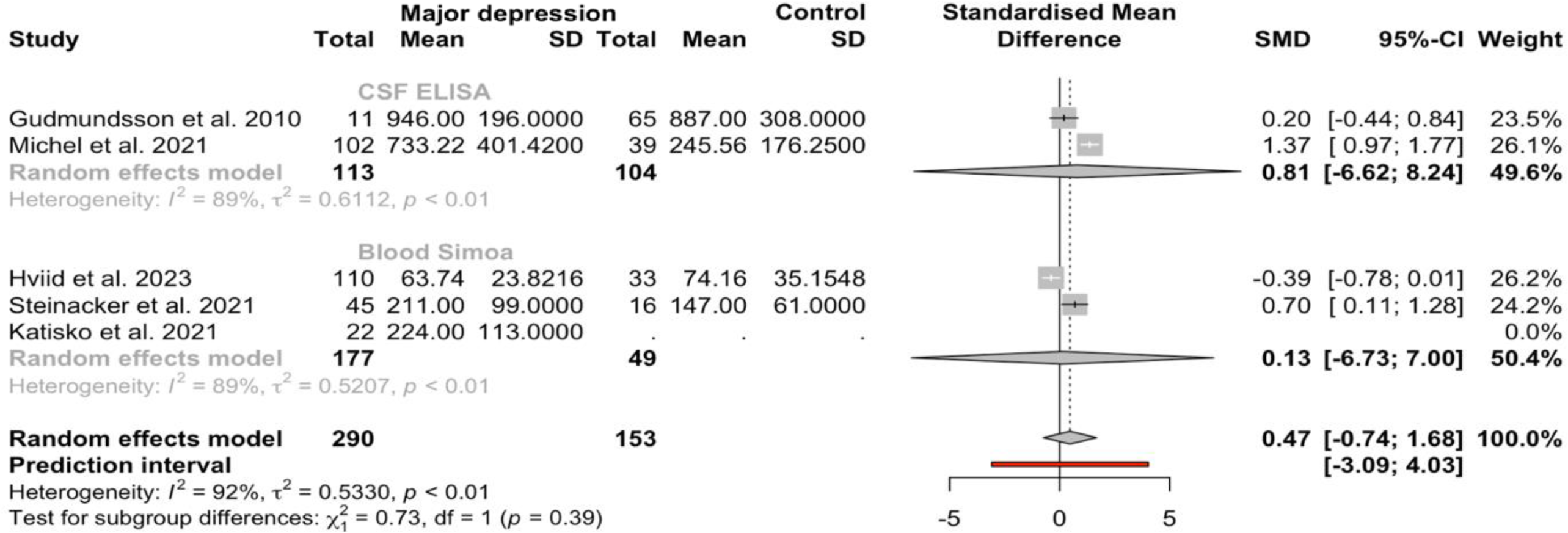
Forest plot of GFAP in major depression

### Bipolar disorder

#### Neurofilament light chain

***All studies (Figure 3a):***The mean concentration of NfL in people with bipolar disorder was significantly higher than healthy control (SMD = 0.58; 95% CI: 0.16, 0.99). However, the prediction interval for this pooled mean difference was wide (PI: -0.67, 1.82) with significant heterogeneity (I^2^ = 84.0%, tau^2^ = 0.24). As Knorr et al’s study (2022) reported both CSF ELISA and plasma Simoa samples, we performed a sensitivity analysis where we excluded the Simoa dataset, which found NfL was still significantly higher in bipolar disorder compared to healthy controls (SMD = 0.62; 95% CI: 0.14, 1.09), There was no asymmetry on the funnel plot (supplementary eFigure 1), and we could not use Egger’s test due to insufficient number of studies.

**Figure 3a.**
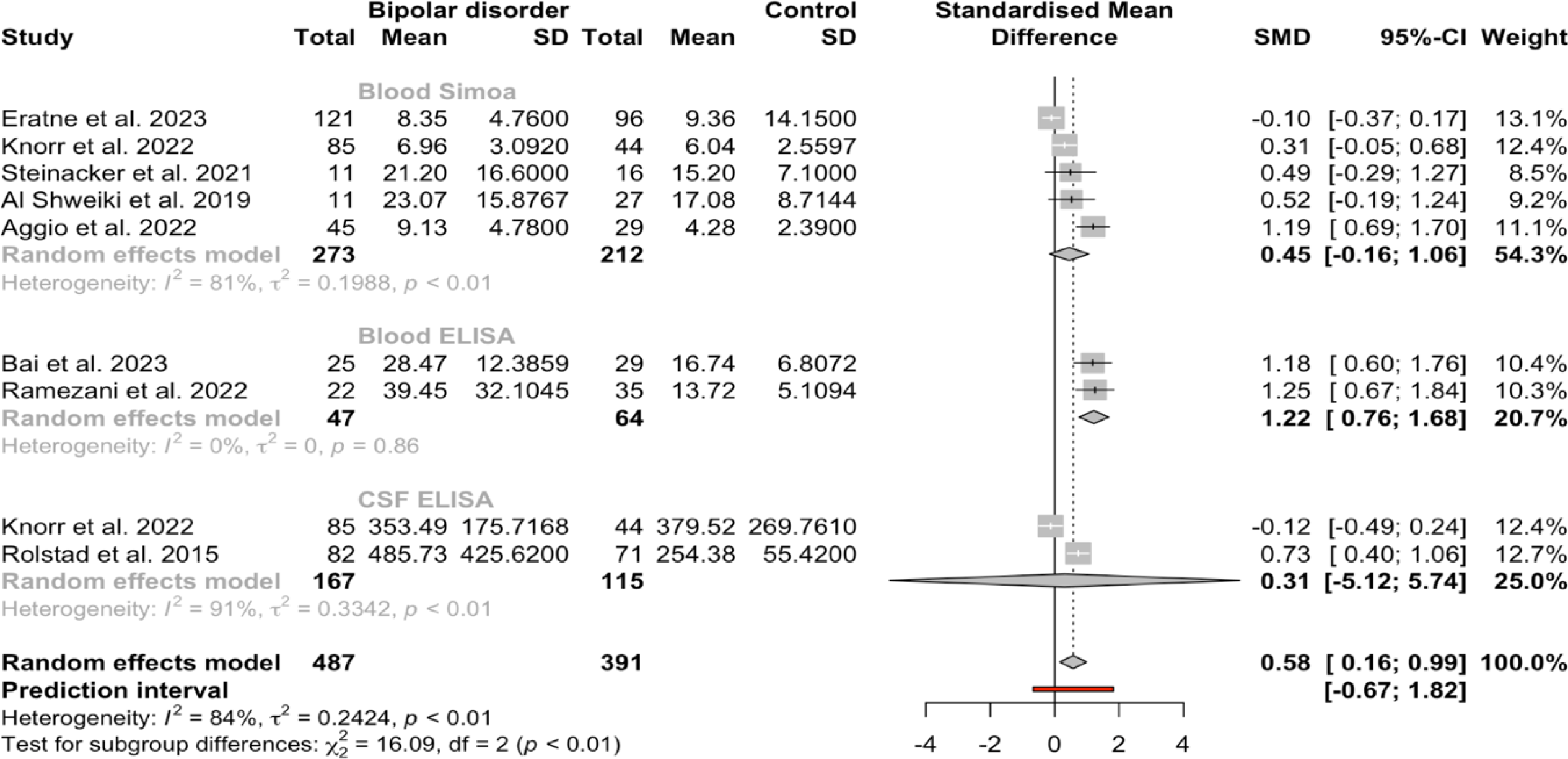
Forest plot of NfL in bipolar disorder (all studies)

*The subgroup analysis model with age-matched studies (Figure 3b)* demonstrated significantly higher NfL in bipolar disorder (SMD = 0.50; 95% CI: 0.06, 0.94; PI: -0.74, 1.74). For the subgroup analysis including only CSF ELISA and blood Simoa studies (Figure 3c), there was no significant difference in the mean concentration of NfL between bipolar disorder and healthy controls (SMD = 0.40; 95% CI: -0.04, 0.85) with a wide prediction interval (PI: -0.80, 1.61).

**Figure 3b.**
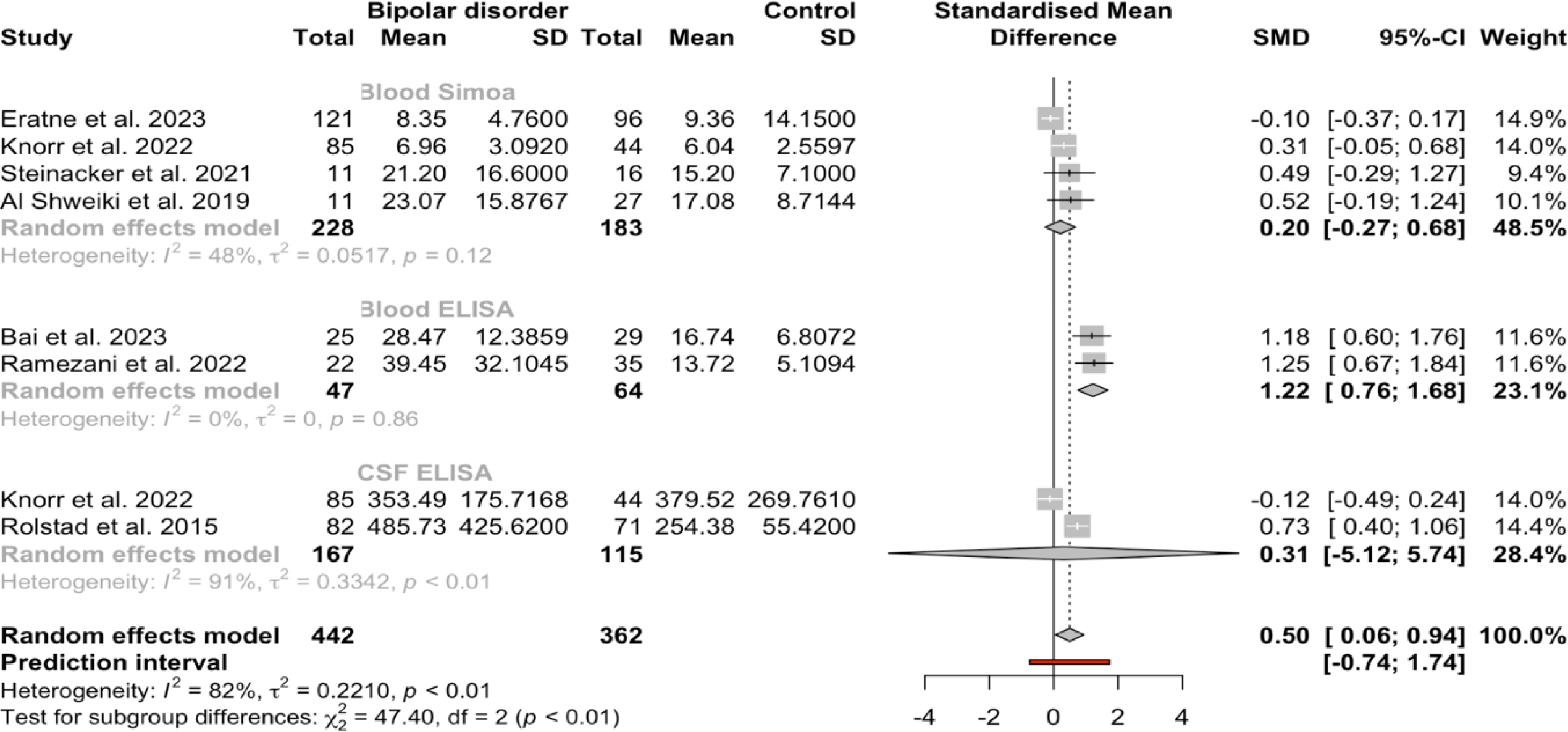
Forest plot of NfL in bipolar disorder (age-matched studies only)

**Figure 3c.**
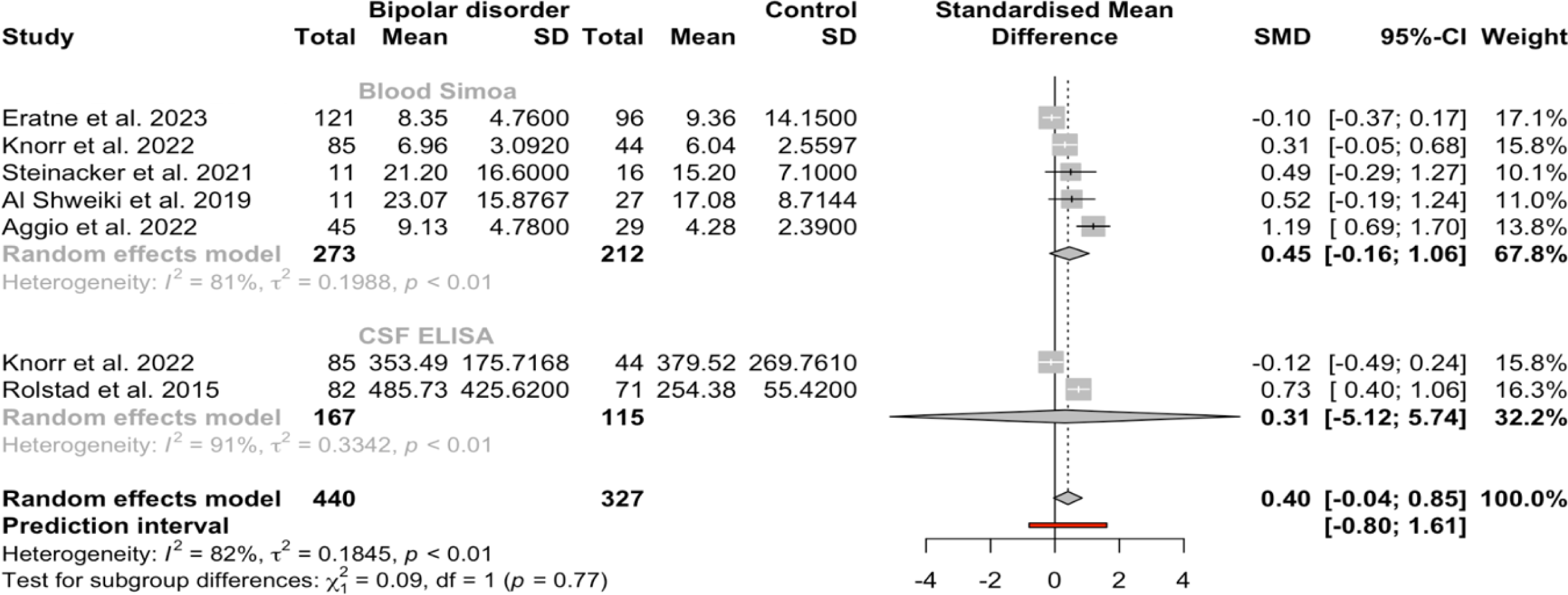
Forest plot of NfL in bipolar disorder (CSF and Simoa only)

Both models were heterogenous (I^2^ = 82.0%, tau^2^ = 0.22 and I^2^ = 81.8%, tau^2^ = 0.18 respectively). The subgroup analysis including six age-matched studies which used CSF ELISA and Simoa also found no significant difference in mean NfL between bipolar disorder and healthy controls. (SMD = 0.27 95% CI: -0.12, 0.65). We performed a subgroup mixed-model mean analysis of blood Simoa studies which showed no difference in NfL concentrations.

Glial fibrillary acidic protein

Only one study (17) studied GFAP in people with bipolar disorder (n=11), which found GFAP concentrations were similar in bipolar disorder compared to healthy controls (125 ± 50 vs 147 ± 61 pg/mL).

## Discussion

Our systematic review found that NfL and GFAP concentrations were not elevated in people with major depression compared to healthy controls. NfL was elevated in people with bipolar disorder, though the difference was no longer significant with subgroup analysis using more sensitive assay kits (CSF ELISA and blood Simoa). We identified no studies examining NfL or GFAP in people with anxiety disorders. To the best of our knowledge, this is the most comprehensive systematic review of these biomarkers in people with primary mood and anxiety disorders.

Our findings align with the existing literature supporting the use of these biomarkers in the differentiation between mood disorders and neurodegenerative disorders (2,5,6). The complex relationship between mood disorders and neurodegenerative disorders, which can present with overlapping symptoms or co-occur, necessitates careful assessment. These findings provide reassurance to clinicians that people with primary mood disorders are unlikely to have very elevated concentrations of NfL and GFAP, as seen in the majority of neurodegenerative disorders where there often is marked elevation (for example, 1.86-fold in Alzheimer’s disease, 3.91-fold in frontotemporal dementia; 44,45).

In individuals with mood disorders, we noted variable instances where NfL concentration was elevated compared to healthy controls, indicating that neuronal injury or neuroinflammation might be present in a specific subset. The limitations of the available data, including detailed clinical and treatment information, prevented us from being able to investigate this further. We plan to conduct a ‘mega-analysis’ by pooling the raw data from the studies included. This approach will enable us to perform statistical adjustments for known covariates including as age, sex and weight/BMI, which were not possible in the meta-analysis. Moreover, identifying specific clusters of symptomatology that correlate with NfL/GFAP may offer valuable insights into the neurodegenerative and neuroinflammatory phenotypes within mood disorders. In turn, opens a new paradigm of diagnosis and treatment for these patients based on biomarker data as well as reported symptoms, in line with the RDOC Framework (56).

Despite the widespread prevalence and burden of anxiety disorders (13), no prior research has explored NfL and GFAP in individuals with primary anxiety disorders. This is an important gap in the literature that this systematic review has identified for the use of NfL and GFAP in clinical practice.

The review process faced several limitations. The evidence was constrained by the quality and heterogeneity of included studies. Controls were not consistently age- and sex-matched (6,18), and some studies lacked control subjects (41,42,51). The limited number of studies and data precluded adequate adjustment for known covariates of NfL and GFAP, such as age, sex, weight/BMI, and renal function (2). This also hindered exploration of factors like mood disorder states (euthymic vs depressed) or effects of psychotropic medications (46). Furthermore, to harmonise the dataset, some of the meta-analyses relied on estimates derived from raw data, which may have introduced potential inaccuracies in the true estimates. Despite our attempts to include as many articles as possible by searching important databases, grey literature, as well as snowballing for other relevant articles, it is possible that some studies may have been inadvertently omitted.

### Conclusions

Our primary analysis indicated that NfL and GFAP levels were not significantly elevated in major depression, while in bipolar disorder, NfL was elevated compared to controls. Subgroup analysis using more sensitive assays revealed no notable differences. These findings suggest that NfL and GFAP elevations in mood disorders are minimal, contrasting with the large elevations in neurodegenerative disorders. This supports the use of these biomarkers in clinical settings to distinguish psychiatric from neurodegenerative causes. The practicality of blood-based biomarker measurements offers valuable diagnostic insights to answer the common clinical question faced by patients, carers and clinicians.

## Data Availability

Data available at reasonable request

## Acknowledgements

We would like to thank Dr Mu-Hong Chen, Dr Leila Simani, and Dr Wen-Yin Chen for their assistance in providing their published data separately to be included in the meta-analysis. We are also grateful for the correspondence from the following researchers who assisted us in clarifying questions about their published data: Dr Anja Fernqvist, Dr Sterre de Boer, Dr Naghmeh Nikkheslat, Prof Carmine Pariante, Dr. Pedro J. Serrano Castro, Dr Johanna Wallensten, Dr. Carol Van Hulle, Dr Mark Miller, Dr Daniel Alcolea, Dr. Suzanne Schindler, and Dr. Andrew Aschenbrenner.

---

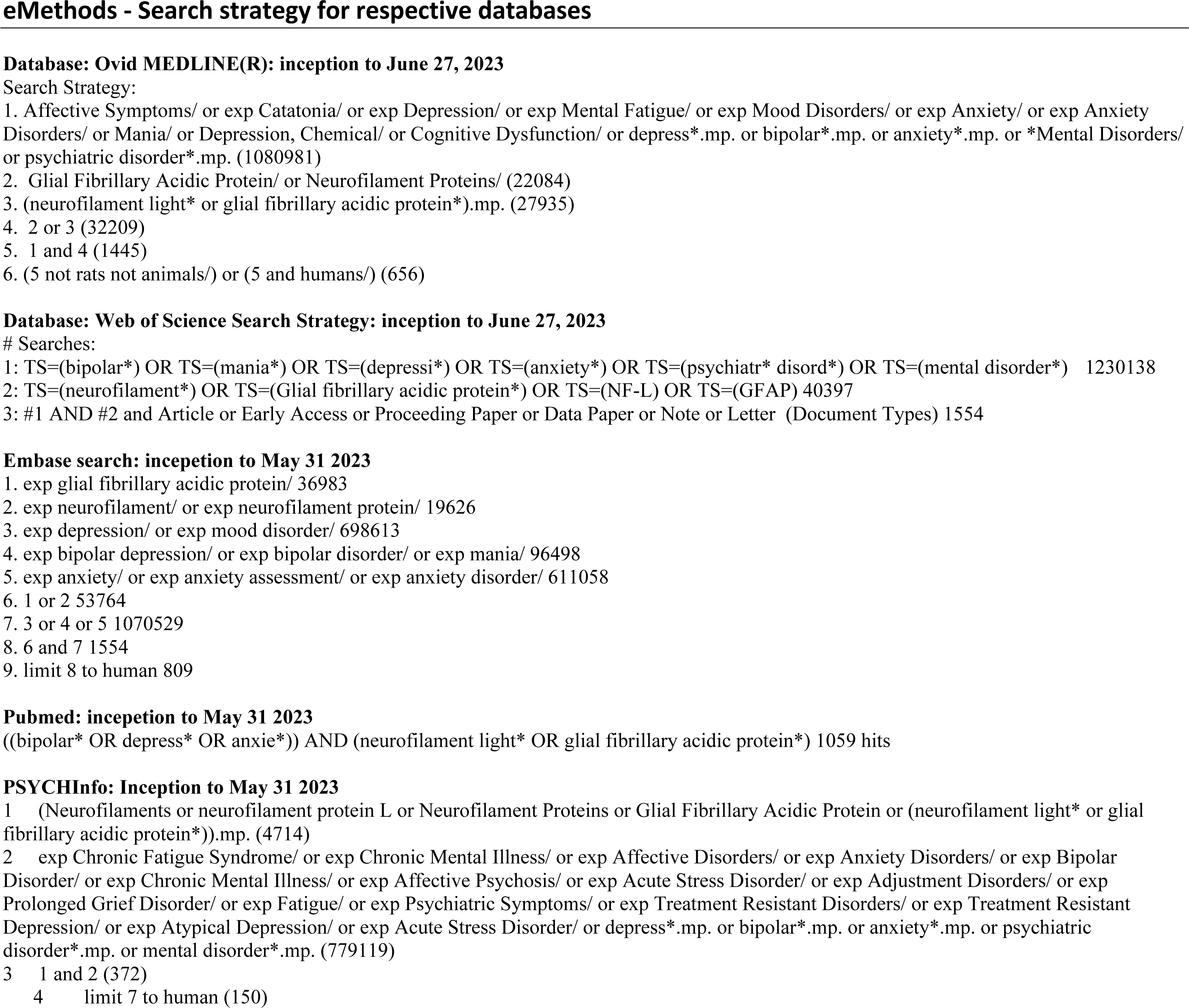

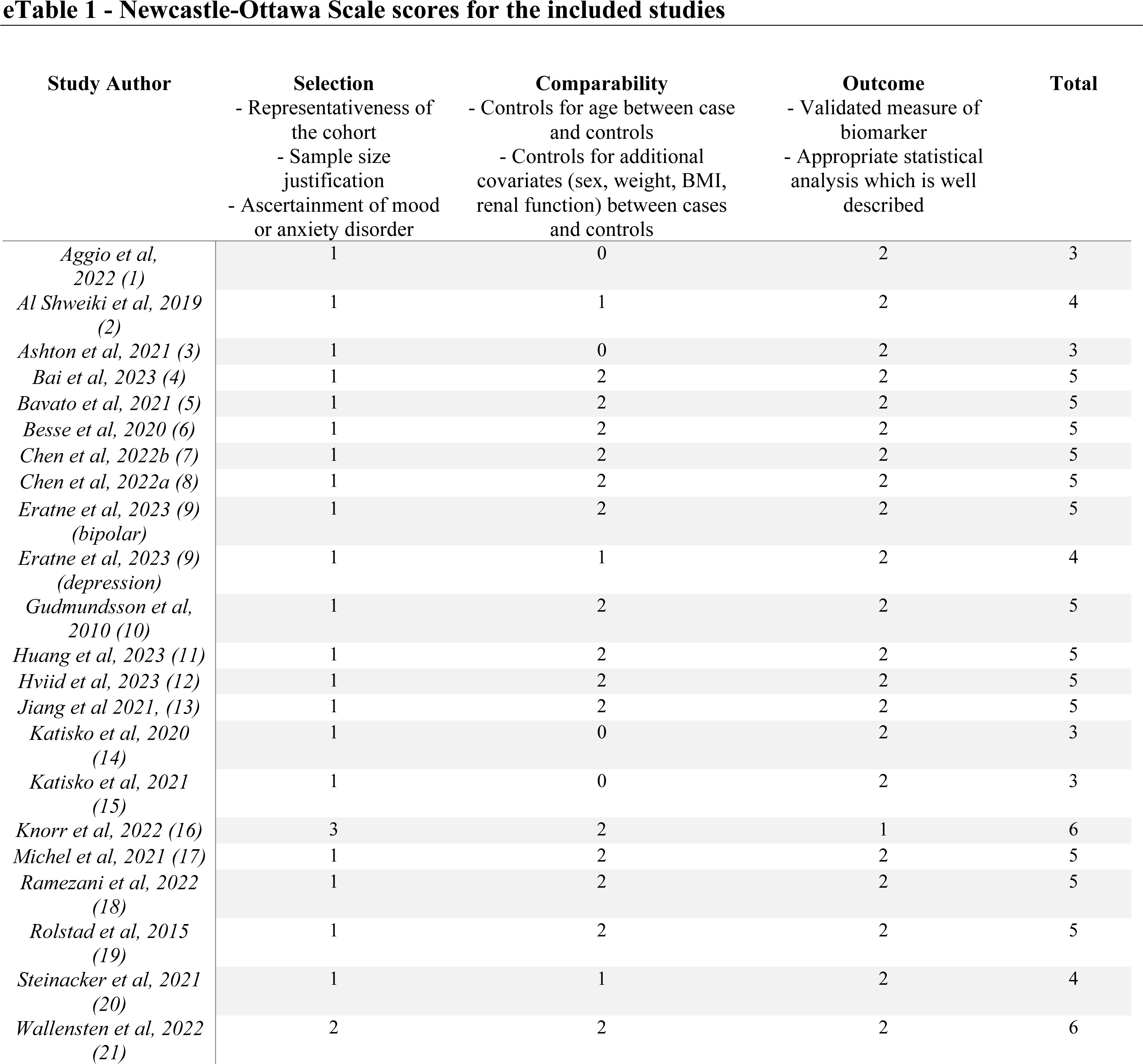

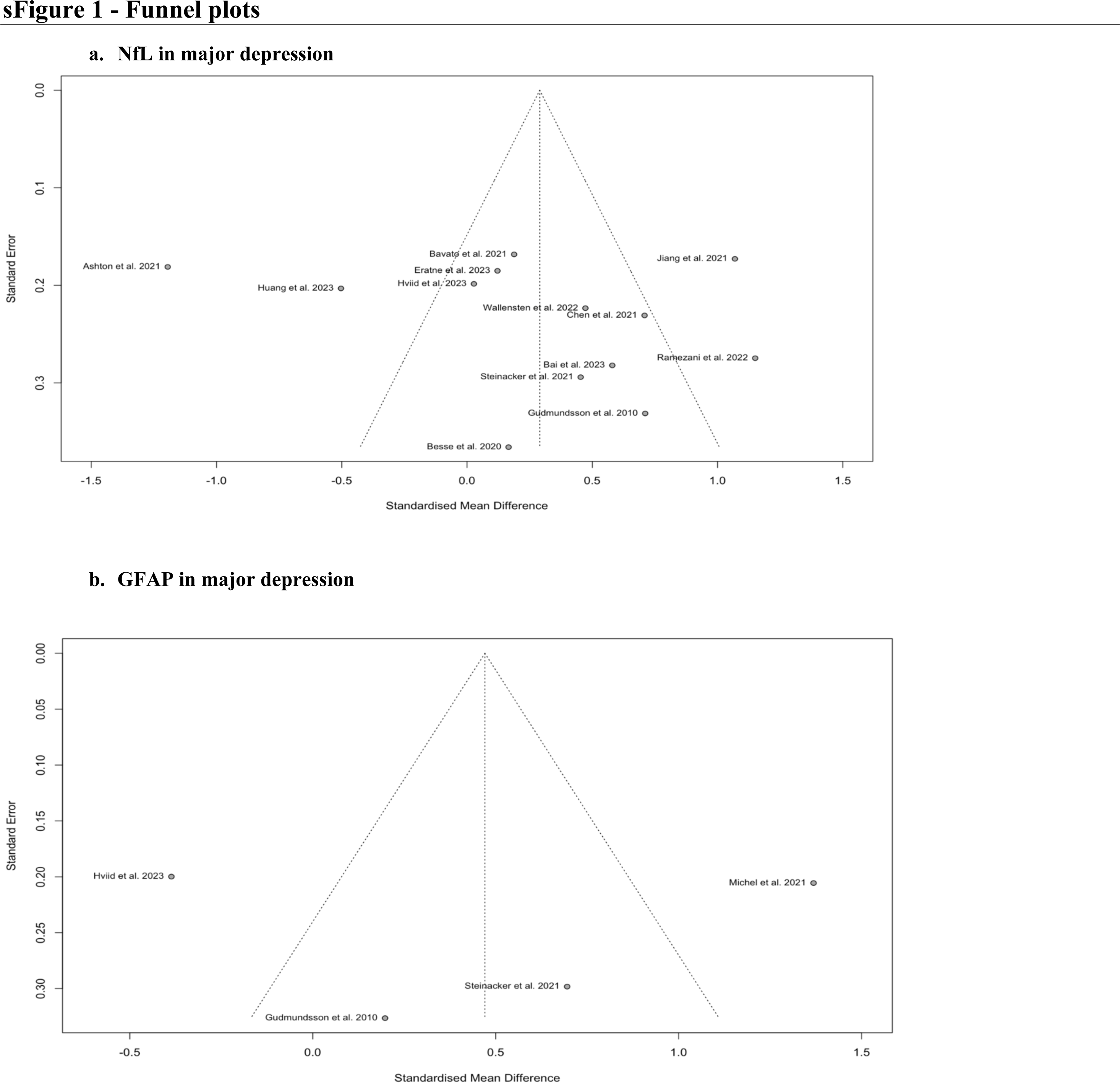
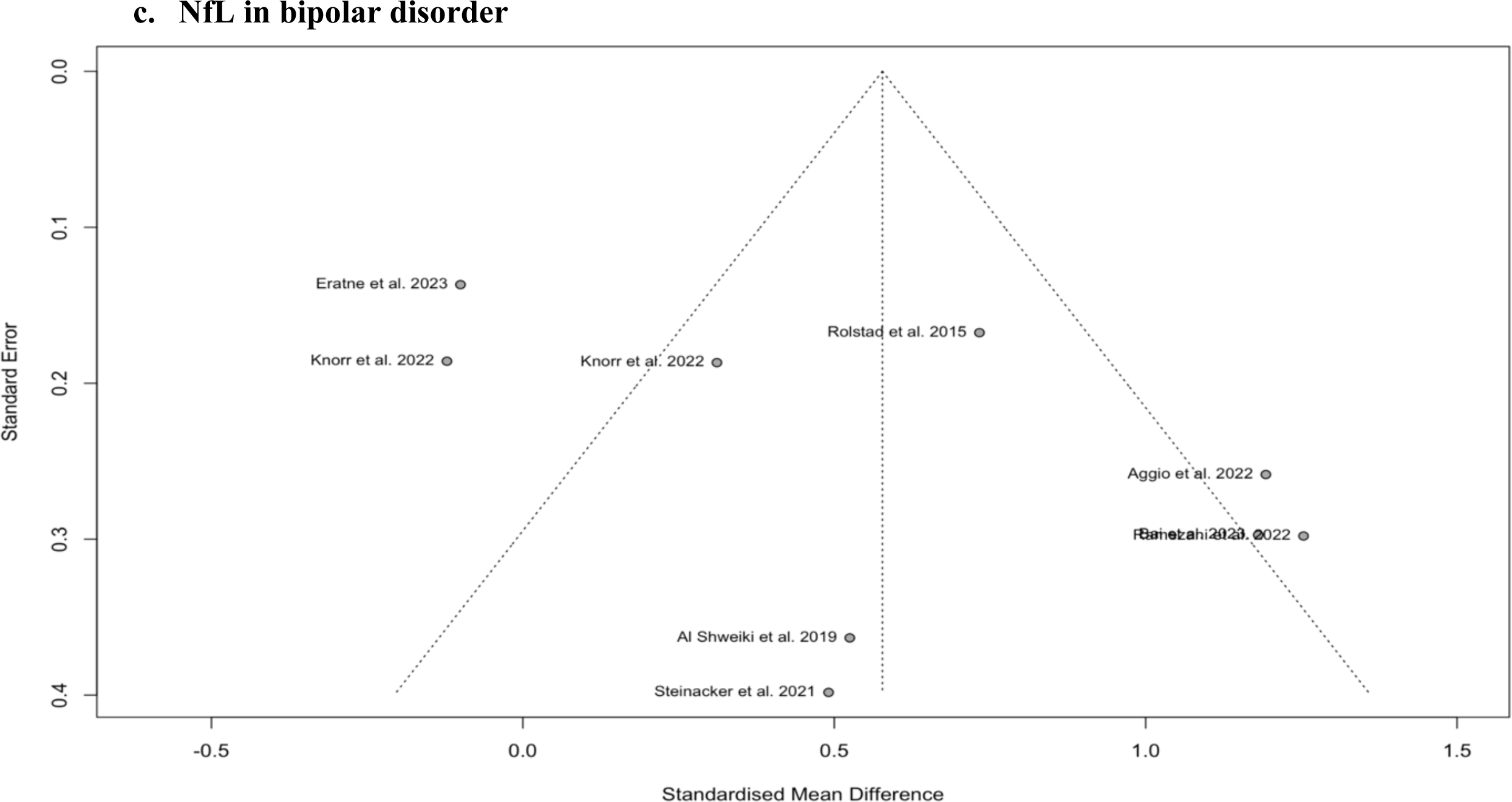

PRISMA 2020 Checklist
| Section and Topic | Item # | Checklist item | Location where item is reported |
| --- | --- | --- | --- |
| **TITLE** |  |  |  |
| Title | 1 | Identify the report as a systematic review. | Pg 1 |
| **ABSTRACT** |  |  |  |
| Abstract | 2 | See the PRISMA 2020 for Abstracts checklist. | Pg 4-5 |
| **INTRODUCTION** |  |  |  |
| Rationale | 3 | Describe the rationale for the review in the context of existing knowledge. | Pg 6/line 89 |
| Objectives | 4 | Provide an explicit statement of the objective(s) or question(s) the review addresses. | Pg 7/line 133 |
| **METHODS** |  |  |  |
| Eligibility criteria | 5 | Specify the inclusion and exclusion criteria for the review and how studies were grouped for the syntheses. | Pg 7/line 144 |
| Information sources | 6 | Specify all databases, registers, websites, organisations, reference lists and other sources searched or consulted to identify studies. Specify the date when each source was last searched or consulted. | Pg 8/line 156 |
| Search strategy | 7 | Present the full search strategies for all databases, registers and websites, including any filters and limits used. | Supplementary |
| Selection process | 8 | Specify the methods used to decide whether a study met the inclusion criteria of the review, including how many reviewers screened each record and each report retrieved, whether they worked independently, and if applicable, details of automation tools used in the process. | Pg 8/line 164 |
| Data collection process | 9 | Specify the methods used to collect data from reports, including how many reviewers collected data from each report, whether they worked independently, any processes for obtaining or confirming data from study investigators, and if applicable, details of automation tools used in the process. | Pg 8/line 171 |
| Data items | 10a | List and define all outcomes for which data were sought. Specify whether all results that were compatible with each outcome domain in each study were sought (e.g. for all measures, time points, analyses), and if not, the methods used to decide which results to collect. | Pg 8/line 171 |
|  | 10b | List and define all other variables for which data were sought (e.g. participant and intervention characteristics, funding sources). Describe any assumptions made about any missing or unclear information. | Pg 8/line 171 |
| Study risk of bias assessment | 11 | Specify the methods used to assess risk of bias in the included studies, including details of the tool(s) used, how many reviewers assessed each study and whether they worked independently, and if applicable, details of automation tools used in the process. | Pg 9/line 185 |
| Effect measures | 12 | Specify for each outcome the effect measure(s) (e.g. risk ratio, mean difference) used in the synthesis or presentation of results. | Pg 8/line 171 |
| Synthesis methods | 13a | Describe the processes used to decide which studies were eligible for each synthesis (e.g. tabulating the study intervention characteristics and comparing against the planned groups for each synthesis (item #5)). | Pg 7/line 186 |
|  | 13b | Describe any methods required to prepare the data for presentation or synthesis, such as handling of missing summary statistics, or data conversions. | Pg 8/line 173 |
|  | 13c | Describe any methods used to tabulate or visually display results of individual studies and syntheses. | Pg 9/line 182 |
|  | 13d | Describe any methods used to synthesize results and provide a rationale for the choice(s). If meta-analysis was performed, describe the model(s), method(s) to identify the presence and extent of statistical heterogeneity, and software package(s) used. | Pg 9/line 182 |
|  | 13e | Describe any methods used to explore possible causes of heterogeneity among study results (e.g. subgroup analysis, meta-regression). | Pg 9/line 205 |
|  | 13f | Describe any sensitivity analyses conducted to assess robustness of the synthesized results. | Pg 9/line 205 |
| Reporting bias assessment | 14 | Describe any methods used to assess risk of bias due to missing results in a synthesis (arising from reporting biases). | Pg 10/line 221 |
| Certainty assessment | 15 | Describe any methods used to assess certainty (or confidence) in the body of evidence for an outcome. | Pg 9/line 201 |

PRISMA 2020 Checklist
| Section and Topic | Item # | Checklist item | Location where item is reported |
| --- | --- | --- | --- |
| **RESULTS** |  |  |  |
| Study selection | 16a | Describe the results of the search and selection process, from the number of records identified in the search to the number of studies included in the review, ideally using a flow diagram. | Figure 1 |
|  | 16b | Cite studies that might appear to meet the inclusion criteria, but which were excluded, and explain why they were excluded. | Pg 10/line 233 |
| Study characteristics | 17 | Cite each included study and present its characteristics. | Table 1-2 |
| Risk of bias in studies | 18 | Present assessments of risk of bias for each included study. | Supplementary eTable 1 |
| Results of individual studies | 19 | For all outcomes, present, for each study: (a) summary statistics for each group (where appropriate) and (b) an effect estimate and its precision (e.g. confidence/credible interval), ideally using structured tables or plots. | Pg 11/line 264<br>Figures 2-3 |
| Results of syntheses | 20a | For each synthesis, briefly summarise the characteristics and risk of bias among contributing studies. | Supplementary eTable 1 |
|  | 20b | Present results of all statistical syntheses conducted. If meta-analysis was done, present for each the summary estimate and its precision (e.g. confidence/credible interval) and measures of statistical heterogeneity. If comparing groups, describe the direction of the effect. | Pg 11/line 263 |
|  | 20c | Present results of all investigations of possible causes of heterogeneity among study results. | Pg 11/line 267 |
|  | 20d | Present results of all sensitivity analyses conducted to assess the robustness of the synthesized results. | Pg 11/line 73 |
| Reporting biases | 21 | Present assessments of risk of bias due to missing results (arising from reporting biases) for each synthesis assessed. | Supplementary eFigure 1 |
| Certainty of evidence | 22 | Present assessments of certainty (or confidence) in the body of evidence for each outcome assessed. | Pg 11/line 268 |
| **DISCUSSION** |  |  |  |
| Discussion | 23a | Provide a general interpretation of the results in the context of other evidence. | Pg 13/line 320 |
|  | 23b | Discuss any limitations of the evidence included in the review. | Pg 14/line 355 |
|  | 23c | Discuss any limitations of the review processes used. | Pg 14/line 360 |
|  | 23d | Discuss implications of the results for practice, policy, and future research. | Pg 15/line 368 |
| **OTHER INFORMATION** |  |  |  |
| Registration and protocol | 24a | Provide registration information for the review, including register name and registration number, or state that the review was not registered. | Pg 7/line 143 |
|  | 24b | Indicate where the review protocol can be accessed, or state that a protocol was not prepared. | Pg 7/line 143 |
|  | 24c | Describe and explain any amendments to information provided at registration or in the protocol. | N/A |
| Support | 25 | Describe sources of financial or non-financial support for the review, and the role of the funders or sponsors in the review. | Pg 2/line 14 |
| Competing interests | 26 | Declare any competing interests of review authors. | Pg 2/line 25 |
| Availability of data, code and other materials | 27 | Report which of the following are publicly available and where they can be found: template data collection forms; data extracted from included studies; data used for all analyses; analytic code; any other materials used in the review. | N/A |

